# Clinician Insights into Effective Components, Delivery Characteristics, and Implementation Strategies of Ambulatory Palliative Care for People with Heart Failure: A Qualitative Analysis

**DOI:** 10.1101/2024.04.08.24305524

**Authors:** Shelli Feder, Lynne Iannone, Dora Lendvai, Yan Zhan, Kathleen Akgün, Mary Ersek, Carol Luhrs, Larry A. Allen, David B. Bekelman, Nathan Goldstein, Dio Kavalieratos

**Author notes:** Corresponding Author: Shelli Feder, PhD, APRN, FNP-C, ACHPN, FPCN, FAHA, Yale School of Nursing PO Box 27399, West Haven, CT 06516.

## Abstract

**Objectives:** To elicit perspectives from specialist palliative care (SPC) and cardiology clinicians on the necessary components, delivery characteristics, and implementation strategies of successful ambulatory SPC for people with heart failure (HF).

**Background:** Palliative care is a recommended component of guideline-directed care for people with HF. However, optimal strategies to implement SPC within ambulatory settings are unknown.

**Methods:** Following a positive deviance frame, we conducted a qualitative study comprising interviews with SPC and cardiology clinicians at Veterans Affairs Medical Centers (VAMCs) with the highest number of ambulatory SPC consultations within the VA system among people with HF from 2021-2022. Clinicians were asked how they provided ambulatory SPC and what they felt were the necessary components, delivery characteristics, and implementation strategies of care delivery. Interviews were analyzed using content analysis.

**Results:** We interviewed 14 SPC clinicians and 9 cardiology clinicians at seven national VAMCs; 43% were physicians 48% were advanced practice registered nurses/physician associates, and 10% were psychologists or social workers. Discussion of goals of care (e.g., prognosis, advance directives) and connecting patients/caregivers to resources (e.g., homecare) were essential components of ambulatory SPC provided at participating facilities. Clinicians preferred and used integrated (i.e., embedded) approaches to SPC delivery, employed standardized patient selection and referral procedures, and formalized procedures for handoffs to and from SPC. Necessary strategies to address barriers to ambulatory SPC implementation included deploying palliative champions, educating non-SPC clinicians on the value of ambulatory SPC for people with HF, and developing ambulatory models through leadership support.

**Conclusions/Implications:** Facilitating the broader adoption of ambulatory SPC may be achieved by prioritizing these mutually valued and necessary features of delivery.

## Introduction

The 2022 American Heart Association/American College of Cardiology clinical practice guidelines for the management of heart failure (HF) strongly recommend palliative care for assistance with symptoms, quality of life, and medical decision-making associated with advanced disease.^1^ However, access to specialist palliative care (SPC) among this population remains markedly low, with less than 20% of people with advanced HF receiving SPC services.^2, 3^ Nationally, SPC is primarily provided to people with HF within inpatient settings. This contrasts with SPC delivery among people with cancer, who have a similar symptom burden,^4^ where 95% of cancer centers in the United States provide ambulatory SPC.^5^ SPC delivered to people with HF primarily within the inpatient setting limits access to those who are acutely ill and reduces opportunities for rapport-building, follow-up, and intervention post-discharge.^6^

The Department of Veterans Affairs (VA) is one of the largest national providers of cardiology services, caring for over 87,000 people with HF annually.^7^ All VA Medical Centers (VAMC) have SPC teams, with services available to any person with serious illness. Rates of SPC consultation among people with HF in the VA are generally higher than non-VA populations;^8^ however, access across the integrated health system varies widely, particularly access to ambulatory SPC.^9^ Interest in developing ambulatory models of SPC for people with HF is growing, including within the VA. However, few successful models exist,^10^ with little practical guidance to support and inform the integration of SPC into ambulatory practice. Little is known about how these models should be structured and the strategies clinicians should use to overcome barriers to implementation. Such data are critical to inform the successful development of ambulatory SPC programs that appropriately address the needs of people with HF while also balancing logistical demands including space, staffing, and a limited SPC workforce.^11^

To address these knowledge gaps, we examined ambulatory SPC delivery among VAMCs that have successfully implemented ambulatory SPC for people with HF. In doing so, we sought to identify the necessary components, delivery characteristics, and implementation strategies of such care. Our study was informed by a positive deviance framework which posits that among a group of entities (e.g., medical centers) some consistently deliver high performance (positive deviants) on a specific outcome using strategies and resources already available among the community of organizations.^12^ Such strategies, once identified, can be implemented to improve performance in the broader community.^13^

## Materials and Methods

### Study Design & Conceptual Framework

We conducted a qualitative descriptive study of seven VAMCs with established ambulatory SPC clinics for people with HF. We report our study methods and findings in line with COREQ qualitative reporting criteria.^14^ The study was approved by Yale University and VA Connecticut Healthcare System Institutional Review Boards. Our study was informed by the *preparation* phase of the modified Multiphase Optimization Strategy (MOST) framework.^15^ The 3-phase MOST framework supports the identification and isolation of effective components of biobehavioral interventions. In the *preparation* phase, potential intervention components are identified, optimized, and evaluated in subsequent phases of research.

### Setting, and Sampling Strategy

Interviews took place between 7/1/2021 and 12/31/2022. VAMCs can vary in terms of bedsize and resources across the system. To address this variation and allow for meaningful comparisons across medical centers, we sampled from VAMCs identified as *high complexity* by the VA. Complexity is a composite variable comprised of several measures including patient volume, number of medical center beds and intensive care units, the number of intensive care unit beds, the availability of specialty services (e.g., cardiology), academic affiliations, and research capabilities.^16^ We sought to identify a sample of VAMCs that consistently provided ambulatory SPC to people with HF. To achieve this, we used data from the VA Corporate Data Warehouse to identify high complexity VAMCs with the highest number of new, completed, ambulatory SPC consultations originating from each VAMC’s cardiology department. From this sample, we calculated the quarterly number of new, completed, consultations for ambulatory SPC from fiscal year 2019 through fiscal year 2021 and averaged this number over the study period. We stratified VAMCs by their quarterly consult average and sampled VAMCs from the top quartile in descending order.

### Sampling Strategy Clinicians

We used purposeful, stratified sampling as our sampling strategy.^17^ Specifically, we emailed SPC team leadership at each VAMC in the top quartile of consults and requested interviews. We also asked SPC clinicians to suggest cardiology program clinicians for recruitment. We aimed to recruit various clinicians from SPC and cardiology programs including physicians, advanced practice registered nurses (APRNs), physician assistants (PAs), and social workers. To participate in interviews, SPC clinicians had to self-report their experience providing ambulatory SPC to people with HF and be employed at the VAMC for at least six months. Cardiology clinicians had to self-report their experience referring people with HF to ambulatory SPC and a minimum of six months of VA employment. We excluded residents and fellows from the sample. Recruitment and data collection continued until data saturation, a point at which transcripts became repetitive and no new codes emerged from the data.^18^

### Interview Guide Development and Procedures

We developed a brief seven-question survey to capture features of the SPC team and the ambulatory clinic. Survey questions asked about SPC team staffing and clinic structures. The survey was sent via email to one SPC clinician from each VAMC prior to interviews. We developed the interview guide using an iterative process and with feedback from cardiology and SPC providers known to our research team. The interview began with study consent, followed by questions about clinicians’ demographic information and work history. Next, the interview guide solicited clinicians’ perspectives on ambulatory SPC grouped into three topics: components and characteristics, informed by the preparation phase of MOST, and implementation strategies (Supplement). Questions also asked about changes to ambulatory models (e.g., use of telehealth) resulting from the COVID-19 pandemic. These data are the focus of a separate forthcoming paper. We piloted the interview guide with two SPC clinicians and one cardiology clinician before implementation. Three study investigators, trained in qualitative interviewing, conducted the semi-structured interviews via telephone or by using Microsoft Teams, a videoconferencing platform. All interviews were audio-recorded and professionally transcribed.

### Data Analysis

All transcripts were imported into ATLAS. ti qualitative software (Scientific Software, Berlin, Germany, version 23.0) for data management, coding, and analysis. We used directed content analysis to compare and contrast codes across interviews. Content analysis is a descriptive qualitative analytic approach in which qualitative data is categorized and then interpreted to generate new ideas or themes.^19^ Coding employed a deductive and inductive process. Coders began coding using the three interview topics derived from our study aims (components, characteristics, implementation) as overarching coding categories. A two-person coding team reviewed the first six transcripts for general meaning, then began building a preliminary code key (i.e., codebook) and applying codes under each coding category. The team then independently applied the code key to the next five transcripts creating new codes as they arose from the data. The team met weekly to review transcripts line-by-line and revise and refine code keys. Disagreements in coding were resolved through group consensus. The team shared code keys periodically with the wider study team for feedback. Transcripts were coded and refinements to the code key continued until data saturation. The team then applied the final code key to the remaining transcripts and created code reports. During two analytic meetings, the study team met to examine code frequencies and analyze code reports identifying key components, delivery characteristics, and strategies for implementation. The team kept memos to track changes in the data analysis procedures. To enhance analytic rigor, preliminary analyses were shared with SPC and cardiology participants for feedback.

## Results

We conducted interviews at seven VAMCs from the Northeast, Mid-Atlantic, South, Midwest, and Mountain West regions of the United States (Table 1). Interview sites had on average 5.2 (standard deviation (SD) 2.8) new ambulatory consultations per study quarter and on average 62.7 new consultations (SD = 34.2) during the study period. Across these sites, the mean number of full-time SPC clinicians was 5.3 (standard deviation (SD) = 2.4). All teams had at least one physician, more than 70% had at least one APRN or PA, all reported having at least one social worker, and 43% reported having a clinical psychologist on the team. All sites offered contact information for the SPC team (e.g., business cards) to patients during their first encounter and 57% had a designated clinician assigned to ambulatory clinics. Forty-three percent of SPC teams reported that they embedded SPC clinics into cardiology programs (i.e., integrated delivery). In this model, SPC clinicians cared for people with HF collaboratively with cardiology. SPC visits were often synchronous with cardiology visits, occurring during joint appointments or after cardiology encounters. The remaining medical centers employed separate (i.e., independent) models of ambulatory SPC where consults were dyssynchronous from cardiology, occurring as distinct visits to separate ambulatory clinics.

**Table 1.** Characteristics of the Specialist Palliative Care Teams.

| **Table 1. Characteristics of Specialist Palliative Care Teams** |  |
| --- | --- |
|  | **Mean (SD)** |
| **Average Quarterly New SPC Ambulatory Consultations per VAMC** | 5.2 (2.8) |
| **Average Total New SPC Ambulatory Consultations per VAMC** | 62.7 (34.2) |
| **Number of clinician members** |  |
| Number of full-time clinicians | 5.3 (2.4) |
| Number of part-time clinicians | 2.4 (1.8) |
| **Number of types of clinicians on the team** |  |
| Physicians | 2.6 (1.7) |
| Nurse practitioners/Physician Associates | 1.9 (1.6) |
| Social workers | 1.3 (0.5) |
| Clinical psychologists | 0.7 (1.1) |
| Chaplain | 0.5 (0.5) |
| **Number of SPC teams with each clinician type, N (%)** |  |
| Physicians | 7 (100) |
| Nurse practitioners/Physician Associates | 5 (71.4) |
| Social workers | 7 (100) |
| Clinical psychologists | 3 (42.9) |
| Chaplain | 3 (42.9) |
| **Ambulatory clinic characteristics** | **N (%)** |
| Provide palliative care team contact information at first visit | 7 (100) |
| Have designated clinicians assigned to ambulatory settings | 4 (57.1) |
| Clinic type |  |
| Embedded (integrated) | 3 (42.9) |
| Separate outpatient* | 4 (57.1) |
| Provide other disease-specific ambulatory palliative care clinics | 3 (42.9) |

IQR = interquartile range, SD = standard deviation, SPC = Specialist Palliative Care
\*Can include general ambulatory or heart-failure specific

Interviews lasted on average 32 minutes (range 15-50). We interviewed 23 clinicians total, 14 SPC and 9 cardiology clinicians (Table 2). The mean age of the sample was 43.9 years (SD =8.3). Thirty-nine percent of the sample were physicians and 48% were APRNs. Clinicians had on average 15.7 years (SD = 6.6) of clinical experience following their terminal degree and almost seven years of experience providing SPC or referring patients with HF to SPC (mean = 6.3 years, SD = 4.9). We describe key components, delivery characteristics, and implementation strategies in the following sections (Figure 1).

**Figure 1:**
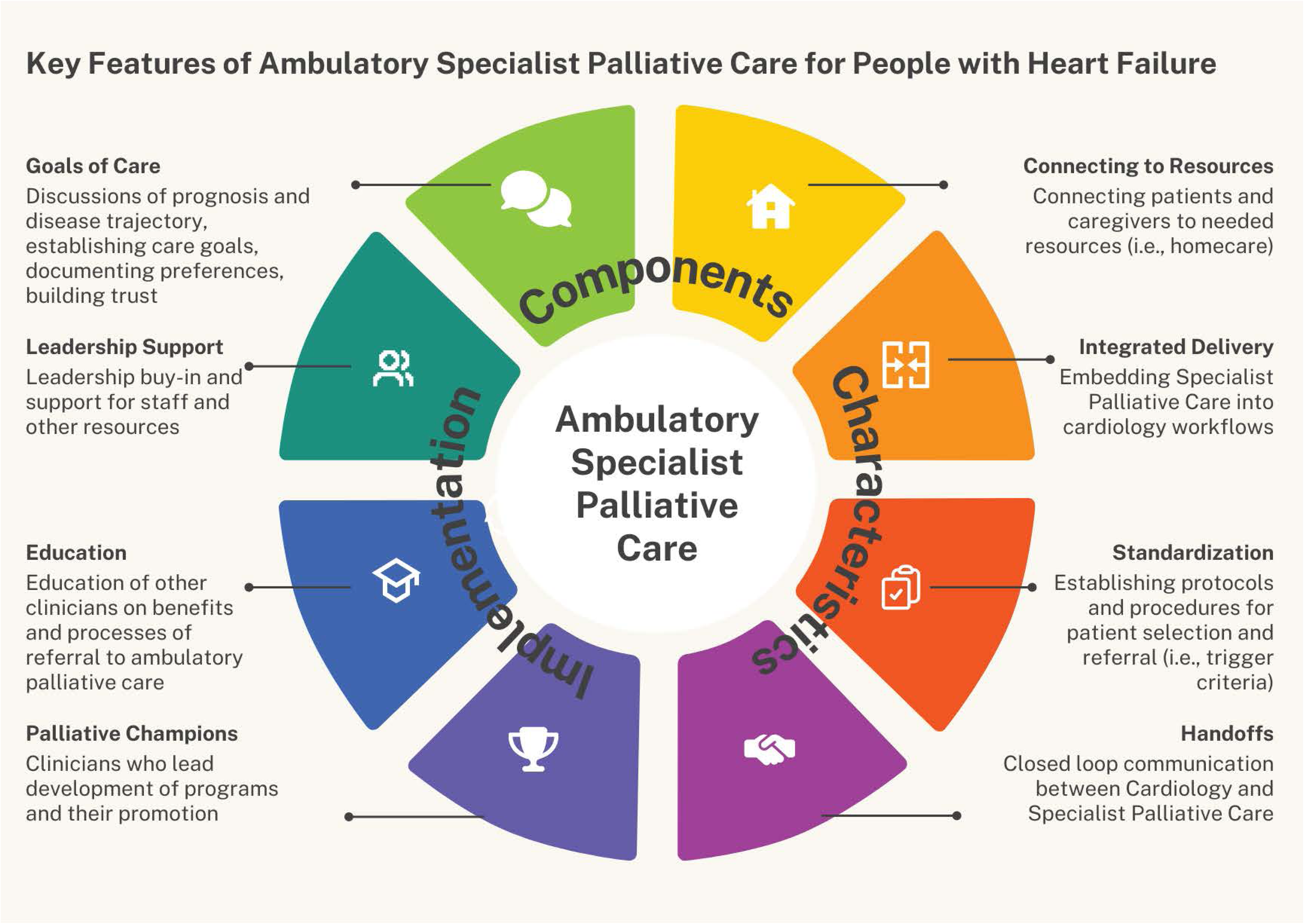
Key Features of Ambulatory Specialist Palliative Care for People with Heart Failure.

**Table 2.** Clinician Characteristics.

| **Table 2. Clinician Characteristics** |  |  |  |
| --- | --- | --- | --- |
| **Variables** | **All Providers**<br>**N=23 (SD ±)** | **SPC Clinicians**<br>**N=14 (SD ±)** | **Cardiology Clinicians**<br>**N=9 (SD ±)** |
| Average age (SD±) | 43.9 (±8.27) | 41.5 (±7.94) | 47.6 (±7.78) |
| Sex |  |  |  |
| Female N (%) | 19 (82.6) | 13 (92.9) | 6 (66.7) |
| Race/Ethnicity, N (%) |  |  |  |
| White/Caucasian | 18 (78.3) | 9 (64.3) | 7 (77.8) |
| Black or African American | 0 (0) | 0 (0) | 0 (0) |
| Asian | 3 (13) | 2 (14.3) | 1 (11.1) |
| Other | 2 (8.7) | 1 (7.1) | 1 (11.1) |
| Hispanic ethnicity | 0 (0) | 0 (0) | 0 (0) |
| Clinician type, N (%) |  |  |  |
| Physician | 10 (43.5) | 4 (28.6) | 4 (50) |
| Advanced Practice Nurse | 10 (43.5) | 5 (35.7) | 4 (50) |
| Physician Associate | 1 (4.3) | 1 (7.1) | 0 (0) |
| Social Worker | 1 (4.3) | 1 (7.1) | 0 (0) |
| Psychologist | 1 (4.3) | 1 (7.1) | 0 (0) |
| Clinical experience, average years (SD ±)<sup>a</sup> | 15.5 (± 5.95) | 16.9 (± 6.72) | 13.4 (± 4.07) |
| SPC experience, average year (SD ±)<sup>b</sup> | 6.53 (±4.93) | 6.8 (± 5.38) | 6.1 (± 4.4) |

SPC = Specialist Palliative Care; SD = Standard deviation
<sup>a</sup> Time from graduation of medical school or other terminal degree program
<sup>b</sup> Time from graduation of medical school or other terminal degree program

### Key Components

Key components of ambulatory SPC were distinct from those of inpatient SPC delivery; the former was an opportunity to provide longitudinal and comprehensive care, while the latter was an opportunity to address often singular, acute problems for those patients often near the immediate end of life. Interventions viewed as essential and delivered within ambulatory SPC clinics for people with HF included discussions of goals of care and connecting patients and caregivers to resources (Table 3; Figure 1). Clinicians varied in their perceived value of symptom management as a necessary component provided during ambulatory SPC encounters.

**Table 3.** Components of Ambulatory SPC Delivery for People with Heart Failure.

|  |  |
| --- | --- |
| Component | Sub-Component & Illustrative Quotes |
| Goals of Care | **Discuss Prognosis & Disease Trajectory**<br>So, I think if they can have a consultation that offers the patient more insight into the nature of their condition then that's successful. (ID #8 – SPC)<br>The most valuable thing I think we offer is [to] have those really, you know, nuanced goals of care conversations. A good family meeting to get everyone on the same page and help them hear the diagnosis and prognosis. (ID #15 – SPC) |
|  | **Establish Care Goals**<br>But in the heart failure population, I would say symptom control is not as dominant as it is in other disease processes. I think—you know, in addition to giving them space and time to talk to me, really advanced care planning is really the most helpful thing with the heart failure population. (ID #2 – SPC)<br>I think that's where we've been able to make the most impact in advocating for patients who are critically ill. If we have some sense of who they are and what their values are but also whether they've kind of imposed any limitations on where they wouldn't want medical interventions because we may be the only person that's having that type of conversation. (ID #4 – SPC)<br>We always talk about goals of care. I think I would probably say the things that we really lean on them, is sort of identifying, sort of the key maybe non-cardiac factors that really affects their care and their outcomes. (ID #6 – Cardiology)<br>I think overall support and communication that the patient and the family knows what the next step is and also that they have a liaison, a person that they can go to if they have needs. (ID #10 – Cardiology) |
|  | **Document Preferences**<br>We discuss their spiritual support, go over, you know, who their decision makers are, if they have an advanced directive, if a licensing treatment note has not been completed, I'll usually complete one during the visit if they know what their wishes are. (ID #11 – SPC)<br>To talk about issues of prognosis and life-sustaining treatment preferences and those kinds of things; we spend a lot of time on that, and I think that's really helpful for both patients and caregivers (ID #19 – SPC) |
|  | **Build Trust**<br>Our outpatient providers see them for quite a while, and it can be - so now you're having a conversation with someone that you know and trust and has helped you manage your symptoms for however long, and so that is hugely beneficial for heart failure patients to then talk about goals of care and lines in the sand and things they would and would not be interested in doing, and if they eventually get recommended for advanced heart failure therapies, if they already knew that provider that's a huge plus. (ID #15 – SPC) |
| Connecting patients to resources | From an outpatient perspective, it's more just coordinating the care for the—the entire care for the patient. (ID #10 – Cardiology)<br>Then the practical support too; like arranging for different services in the home is really helpful for caregivers as our patients become more debilitated and require more care. (ID #19 – SPC) |
| Symptom management (physical and psychological) | **Key Component**<br>I think the palliative care people feel a little more comfortable dealing with some of the medications that are needed for people with heart failure that they have symptoms that are resistant to it or failing treatment. (ID #14 – SPC)<br>I think PC is very beneficial for symptom management kind of over the course of the disease and through some of the exacerbations. (ID #19 – SPC)<br>**Not a Key Component**<br>The patient needs to discuss goal of care. We tend not to think about just for symptomatic or spiritual support. We tend not to think about referring only for that. (ID #3 – Cardiology)<br>I think symptom management is actually simpler than helping elucidate a person's goals of care. So, I think a heart failure patient where you know that they're gonna end up back in the intensive care unit at some point trying to get a sense of what their wishes and values are about life sustaining treatments is probably even more important than symptom management. (ID #4 – SPC)<br>An inappropriate referral might be a patient who is being referred for say symptomatic management of their dyspnea, but their dyspnea is completely related to their fluid overload. So the intervention really would be titration of you know their diuretics which is not something that we're gonna step in to do if they've got a cardiology team. (ID #16 – Cardiology) |

SPC = Specialist Palliative Care

#### Goals of Care

Goals of care was a broad term used by both SPC and cardiology clinicians to refer to several related actions by the SPC team: discussions of prognosis and disease trajectory, establishing care goals, documenting preferences, and building trust. Conversations about goals of care that occurred during ambulatory visits encompassed discussions of prognosis and disease trajectory to “achieve a better understanding of [a patient’s] prognosis and what to expect (Cardiology clinician).” These conversations often involved discussions about preferences and care goals for advanced therapies. Topics included the use and deactivation of implantable cardiac defibrillators (ICD) and intravenous inotropes, among others. A cardiology clinician remarked, “You know, did they [SPC] discuss about ICD deactivation, discuss about patient’s understanding of what happened when an ICD deployed.” Clinicians viewed goals of care conversations as opportunities to document surrogate decision-makers and complete advance directives. These were not singular discussions; rather clinicians conducted multiple goals of care discussions over time.

Connecting to resources involved educating and linking patients with HF and their caregivers to appropriate resources including homecare, transportation services, and hospice. This task was often undertaken by the SPC team’s social worker. SPC providers connected patients and their caregivers with resources that addressed immediate needs, but also provided anticipatory guidance of resources available to them in the future. An SPC social worker reported, “I’m talking about services too but not just ones that they need immediately but just saying down the road you might need this type of support.”

For many clinicians, symptom management was not a key component of ambulatory for people with HF. “I think having the presence of palliative care really helps to answer questions and to have the discussion,” reported an SPC clinician, “I don’t do a ton in the way of aggressive symptom management. Really no symptom management.” Cardiology and SPC clinicians often felt that most symptoms could be addressed by guideline-concordant cardiology care. “When I think of oncology palliative care, there’s a lot of symptom control that they need,” a cardiologist reported, “When it comes to HF palliative care, one of the things that I do medically is symptom control.” Others valued SPC’s involvement for symptoms resistant to treatment or for those patients with symptoms near the end of life. “If it’s a HF patient with like imminent death and we’re gonna do inpatient hospice for them, we’ll get consulted and we’ll manage symptoms more for those patients, but that’s not a ton of our consults (SPC clinician).”

### Key Characteristics

Key delivery characteristics implemented within ambulatory SPC clinics for people with HF included the use of integrated delivery models, standardized patient selection and referral procedures, and standardized procedures for handoffs to and from SPC (i.e., setting up and communicating the referral) (Table 4; Figure 1).

**Table 4.**
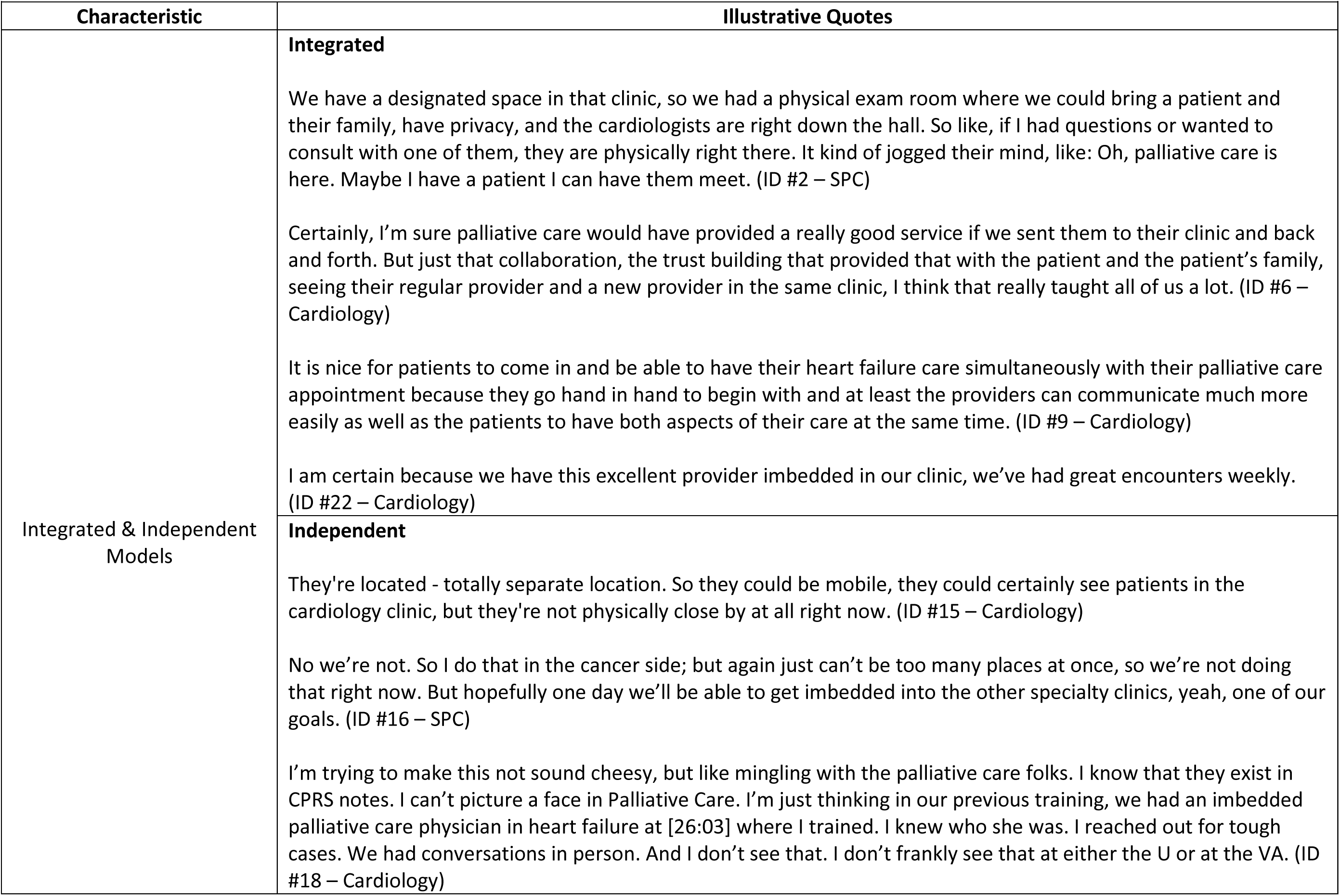
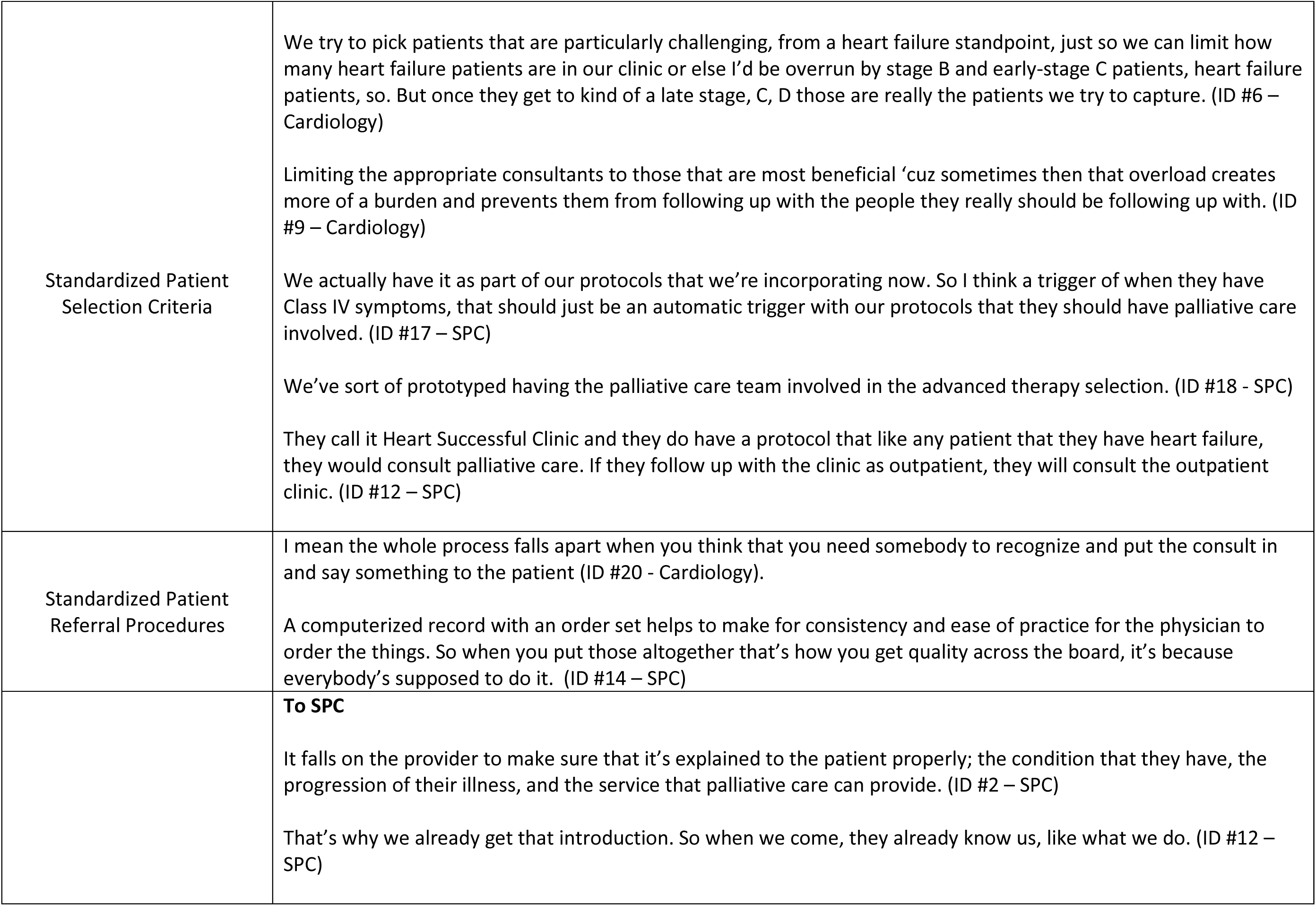
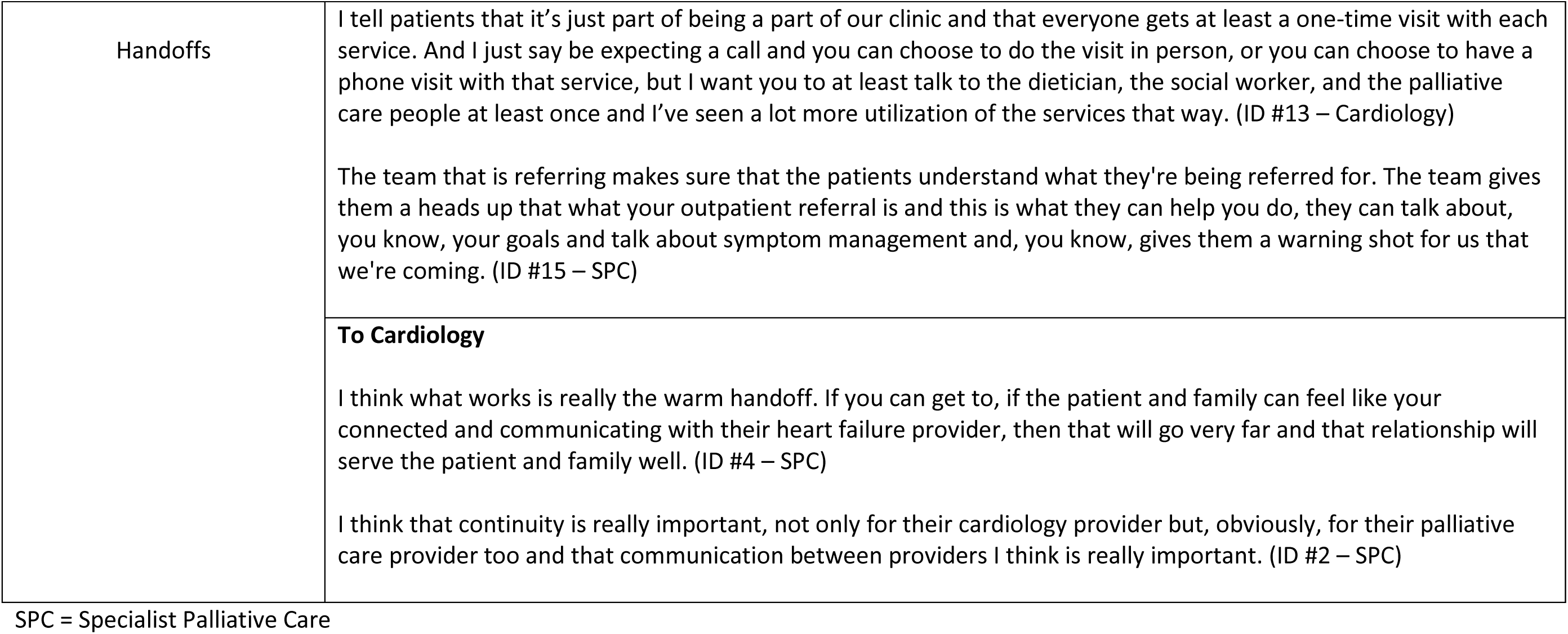
Key Characteristics of Ambulatory SPC for People with Heart Failure.

Integrated models of ambulatory SPC delivery were preferred by most clinicians, even among those from VAMCs with independent ambulatory clinics. Cited benefits of these models included facilitating close collaboration between cardiology and SPC clinicians, building trust, increasing referrals, and facilitating seamless integrated care. “It is nice for patients to come in and be able to have their HF care simultaneously with their palliative care appointment,” an SPC clinician stated, “They go hand in hand to begin with.” A cardiologist reported, “I learned so much from the collaborative part of the clinic. I think I would not have learned as much, and my [APRNS] would not have learned as much, had we not started that way.”

Clinicians preferred and utilized standardized processes for referring patients with HF to ambulatory SPC. Typically, these processes helped to address clinician difficulties with accurate prognostication by using set criteria and procedures to monitor which patients were referred to SPC and when. A cardiology clinician remarked “If we just run around saying, ‘oh do you think this person will be alive in six months or do you think this person needs palliative care?’ The average physician is gonna say, ‘Well I don’t know.’ So it’s better to make it a protocol.” Standardized processes reduced clinician uncertainty and streamlined ordering procedures, saving time during busy clinics. A cardiology clinician stated, “I mean the whole process falls apart when you think that you need somebody to recognize and put in the consult and to limit the appropriate consultations to those that are most beneficial.”

Clinicians used several criteria in referring people with HF to ambulatory SPC including demographic and clinical characteristics (e.g., age, cardiac function), symptoms, functional status (e.g., New York Heart Functional Class), prognosis, and healthcare utilization. Consistency and standardization were key in ensuring quality referrals. Several sites employed consultation triggers and standardized order sets to refer people with HF to ambulatory SPC clinics. Electronic health record dashboards were viewed as a viable way to monitor patients and referrals, and "to make it a successful protocol then you monitor and reward the people who do it (Cardiology clinician).” However, dashboards were not yet employed at the seven VAMCs included in the sample.

Ambulatory SPC encounters began and ended with handoffs; specific processes for setting up the referral and closed-loop communication among SPC and cardiology clinicians about the encounter. Setting up the referral involved conversations between cardiology clinicians and the patient and family about the reason for the referral and an overview of potential services provided by SPC. Phrasing referral to ambulatory SPC as a part of standard HF care “a part of our clinic that every patient receives (Cardiology clinician)” set expectations that ambulatory SPC was essential to high-quality HF care. Handoffs also included SPC clinicians reporting back to their cardiology colleagues after encounters. Integrated ambulatory SPC facilitated this communication by co-locating clinicians within close proximity, facilitating spontaneous conversations and other opportunities for engagement.

### Key Strategies for Implementation

Three common strategies used to implement ambulatory SPC for people with HF included identifying champions, educating cardiology clinicians about the value of ambulatory SPC for people with HF, and identifying and maintaining leadership support (Table 5; Figure 1).

**Table 5.** Key Implementation Strategies of Ambulatory SPC Delivery for People with Heart Failure.

|  |  |
| --- | --- |
| Characteristic | Illustrative Quote |
| Champion | We definitely have a couple champions down there. And then some people who are less familiar with us. (ID #1 – SPC)<br>Being able to attend the evaluation meetings and offer opinions; and be a face that providers know. I think the program, I think the relationship really did strengthen when we had that person. It gave the cardiac team a lot of faith in us too once they kind of knew us and knew what. (ID #16 – SPC) |
| Knowledge/Education | We now present at every new hospital orientation so that everybody's that coming in knows that our team exists somewhere or are happy to be available in any of the setting, you know, as the newer tiers of trainings come out and become attendings and then in more full time positions they're getting a lot more palliative care education at the granular medical school level and I think that will make a difference. (ID #4 – SPC)<br>It's almost the ownership of that ordering physician that they believe in the service as well. (ID #11 – SPC)<br>The education part is very key point to the program because, I mean, we only can accept it if we are well aware of what it is. So I mean for like education to the provider, education to our patient, our community so they aware of us so that way they can accept us more and more. (ID #12 – SPC) |
| Leadership Support | Having someone at an upper executive leadership level who's worked very closely with our team, very directly with pretty great outcomes for patients I think is good because we can pull upon that experience and say, listen, we're still having deaths in the hospital with this diagnosis. (ID #4 – SPC)<br>I think in terms of the Medical Center's responsibility, to ensure that the service is known to the providers that work there so that they can utilize it adequately is probably the most important thing (ID #9 – Cardiology)<br>Our leadership are very supportive of our specialty. They provide us opportunity to share our service and our specialty with a lot of other interdisciplinary teams. (ID #12 – SPC) |

SPC = Specialist Palliative Care

Champions were cardiology and/or SPC clinicians who supported the development and sustainment of the ambulatory SPC clinic. All sites mentioned specific clinicians who engaged in this role and often identified two champions, one from cardiology and one from SPC who engaged collaboratively. Champions were often tasked with defining the initial scope and practice of the clinic, the model for delivery (e.g., integrated, independent), and other clinic structures and processes. Champions also promoted the clinic and encouraged referrals. An SPC clinician discussed why this role at her VAMC was successful by commenting, “being able to attend the evaluation meetings and offer opinions; and be a face that providers know. It gave the cardiac team a lot of faith in us too once they kind of knew us.”

SPC clinicians frequently employed strategies to educate other clinicians about the benefits of ambulatory SPC for people with HF. Education took many forms including in-services, the creation and distribution of educational materials, and point-of-care educational encounters. Outreach was viewed as essential, so that cardiology providers knew of the ambulatory SPC clinic, when to refer, and how to do so. Collaborative educational sessions involving both cardiology and SPC clinicians boosted these efforts. Clinicians also obtained leadership support to incentivize cardiology and SPC clinicians to act in clinic development and assisted in obtaining sufficient staff for the clinic. A cardiologist reported, “So that requires some incentive or stimulus that oftentimes comes from a leadership aspect in talking with a team and reminding them of the importance and availability of this.”

## Discussion

To our knowledge, this is the first study to document the real-world implementation of ambulatory SPC specifically for people with HF across a national sample of medical centers and identify clinicians’ perspectives of the necessary features of these programs. In this study, ambulatory SPC programs engaged people with HF and their families in conversations around goals of care, connected them to resources, and often focused less on symptom management. These programs employed both integrated and independent models of delivery, though clinicians preferred integrated models. In many cases, cardiology and SPC clinicians standardized referral criteria and established procedures to allow for close communication across specialties. Common strategies for implementation included the employment of ambulatory SPC champions who promoted the clinic to other providers and medical center leadership, providing education about the value of SPC to non-SPC providers, and obtaining and sustaining leadership support.

Most clinicians in this study viewed discussions of goals of care as a necessary feature of ambulatory SPC for people with HF. Broadly, these tasks encompassed aspects of serious illness conversations that are part of the HF “care planning umbrella” a term used to describe health communication across the illness trajectory.^20^ It is perhaps not surprising that cardiology clinicians looked to SPC providers in ambulatory settings to provide this aspect of the communication continuum. Cardiology clinicians often report barriers to conducting conversations about goals of care, including discomfort in having these conversations, and a limited amount of training and time to conduct such conversations in busy clinics, among others.^21^

Interestingly, SPC and cardiology clinicians frequently viewed symptom management as a task best addressed by cardiology, unless symptoms were attributable to the end of life. This finding is in stark contrast with oncology, where symptom management is a key pillar of SPC delivery. Clinician sentiments likely reflect evidence that adherence to clinical practice guidelines improves cardiac function and congestion, which in turn reduces symptom burden among people with HF.^1^ However, we did not ask clinicians about the specific symptoms they did or did not address in clinic and some symptoms, when present, may be more readily addressed by SPC.

Our study has significant implications for clinicians, administrators, and healthcare systems wishing to implement ambulatory SPC for their HF populations. First, people with HF have unique palliative needs reflective of their disease course and treatment options. Ambulatory SPC clinics should be developed to address these unique needs; this may impact how these clinics are staffed and even how care is delivered. For example, ambulatory clinics could be designed to focus on aspects of serious illness conversations and care coordination. Visits for advance directive completion or care coordination could be led by palliative social workers or by non-specialist nurses and social workers trained in palliative principles. ^22^ SPC physicians and advance practice providers could be involved in these visits only when necessary, such as visits related to complex decision-making or symptom management. Such a tailored approach could help to increase access to palliative care and alleviate strained SPC clinicians who often face workforce shortages and increasing workloads.^23^

Our findings also highlight key considerations for clinic design and implementation. Clinicians in our study preferred integrated or embedded co-management models, also called collaborative care models,^24^ of ambulatory SPC over independent ones. This may reflect the unique perspectives of VA clinicians practicing within VA ecosystems. However, others have also identified several benefits of integrated ambulatory SPC delivery outside the VA, including increased collaboration and communication among specialty and non-specialty providers and an increased presence of the SPC team resulting in increased referrals.^25^ The ADAPT trial is a recent successful example of a collaborative, integrated approach.^22^ In this trial a nurse and social worker trained in palliative principles provided telehealth palliative care to people with Chronic Obstructive Pulmonary Disease, HF, and Interstitial Lung Disease. The telehealth team met weekly with primary care, a palliative care physician, and as needed, a pulmonologist, and cardiologist to coordinate patient care. The intervention resulted in a significant increase in patient-reported quality of life relative to usual care among the intervention group.

This study has several limitations. With our approach focusing on VAMC sites with high rates of SPC, we elicited perspectives from a relatively narrow group of clinicians from similarly resourced settings. Therefore, while we reached data saturation among clinicians surveyed, important perspectives on SPC may have been missed. We did not interview chaplains, as they function as part of a separate service within the VA system. Finally, we conducted this study during the later stages of the COVID-19 pandemic which altered care models, in some cases significantly, as teams moved to telehealth.^26^ Our findings reflect these changes, and the evolution of ambulatory SPC that occurred during this period.

## Conclusion

Clinicians across seven VA medical centers with established ambulatory SPC clinics identified several necessary components, delivery characteristics, and strategies for implementation for people with HF. Goals of care discussions, care coordination, integrated delivery, standardized patient selection and referral procedures, and formalized handoffs and communication were key and necessary components and characteristics delivered to people with HF in these ambulatory settings. Clinician’s attitudes towards symptom management as a necessary component provided during ambulatory SPC varied. Implementation included deploying palliative champions within cardiology services, educating non-SPC clinicians on the value of ambulatory SPC for people with HF, and developing ambulatory models through leadership support. Facilitating the broader adoption of ambulatory SPC may be achieved by prioritizing the mutually valued and necessary features of delivery.

### Clinical Perspectives

In this qualitative study, SPC and cardiology clinicians across seven VA medical centers with established ambulatory SPC clinics identified necessary components and delivery characteristics of ambulatory SPC for people with HF and strategies for implementation. Prioritizing these mutually valued and necessary features of ambulatory SPC delivery may facilitate the adoption of ambulatory SPC more broadly across care settings and healthcare systems.

## Data Availability

The availability of data from this manuscript is in accordance with the West Haven Department of Veterans Affairs Institutional Review Board and VA research polices.

## Acknowledgments

None

## Abbreviations

HF: Heart Failure
VA: Department of Veterans Affairs
VAMC: VA Medical Center
SPC: Specialist Palliative Care
ICD: Implantable Cardiac Defibrillators
APRN: Advanced Practice Registered Nurse
PA: Physician Assistant

---

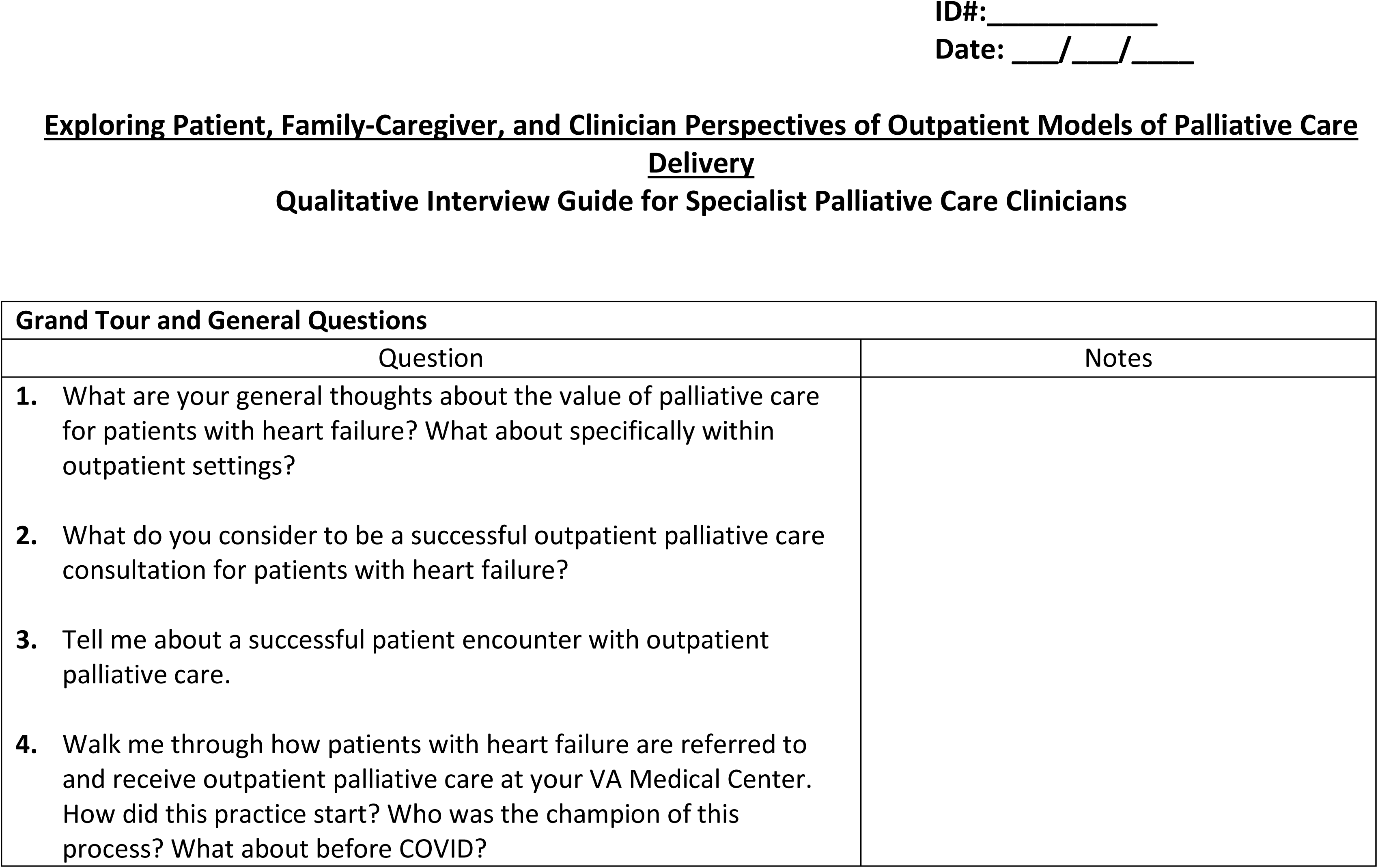
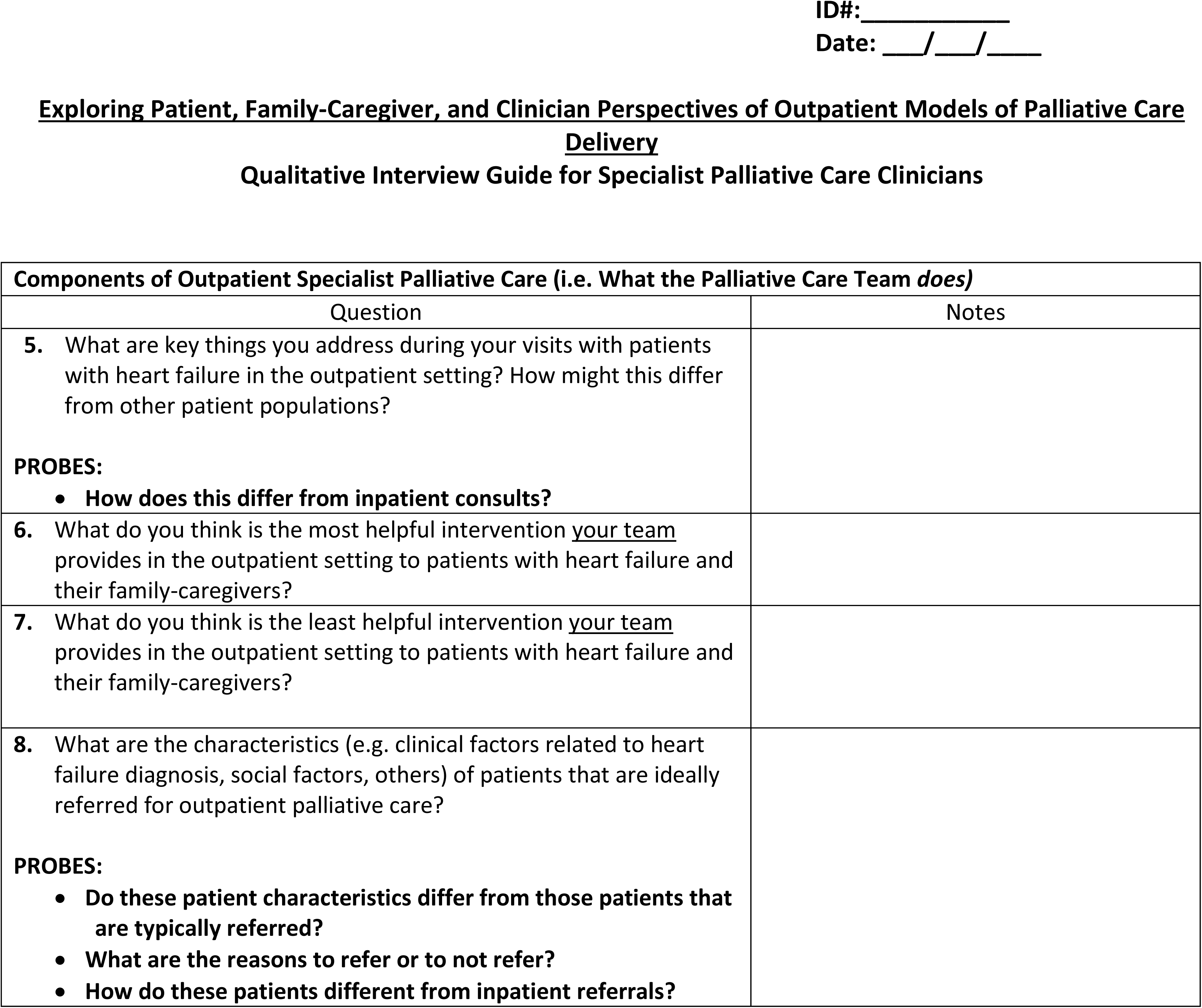
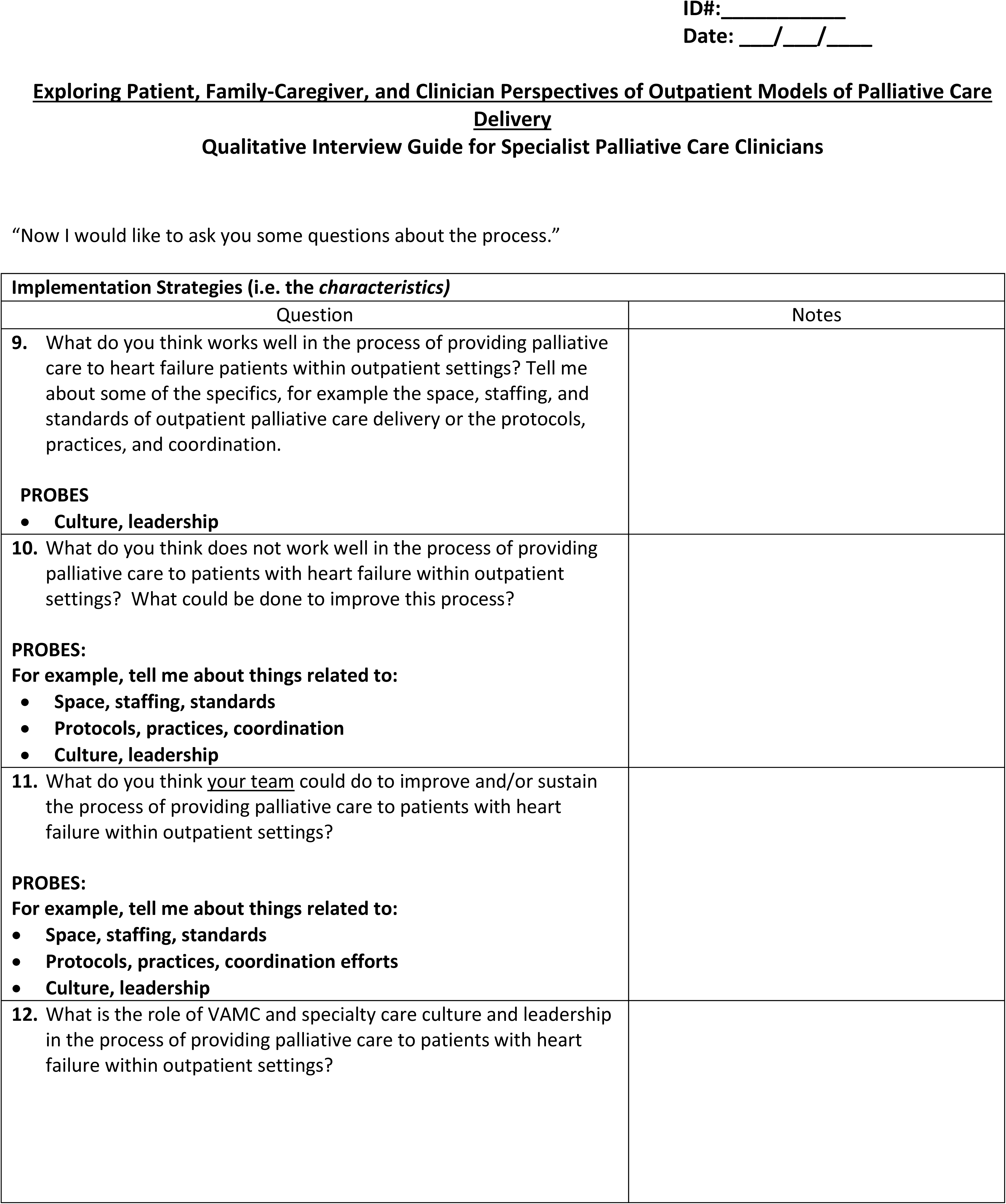
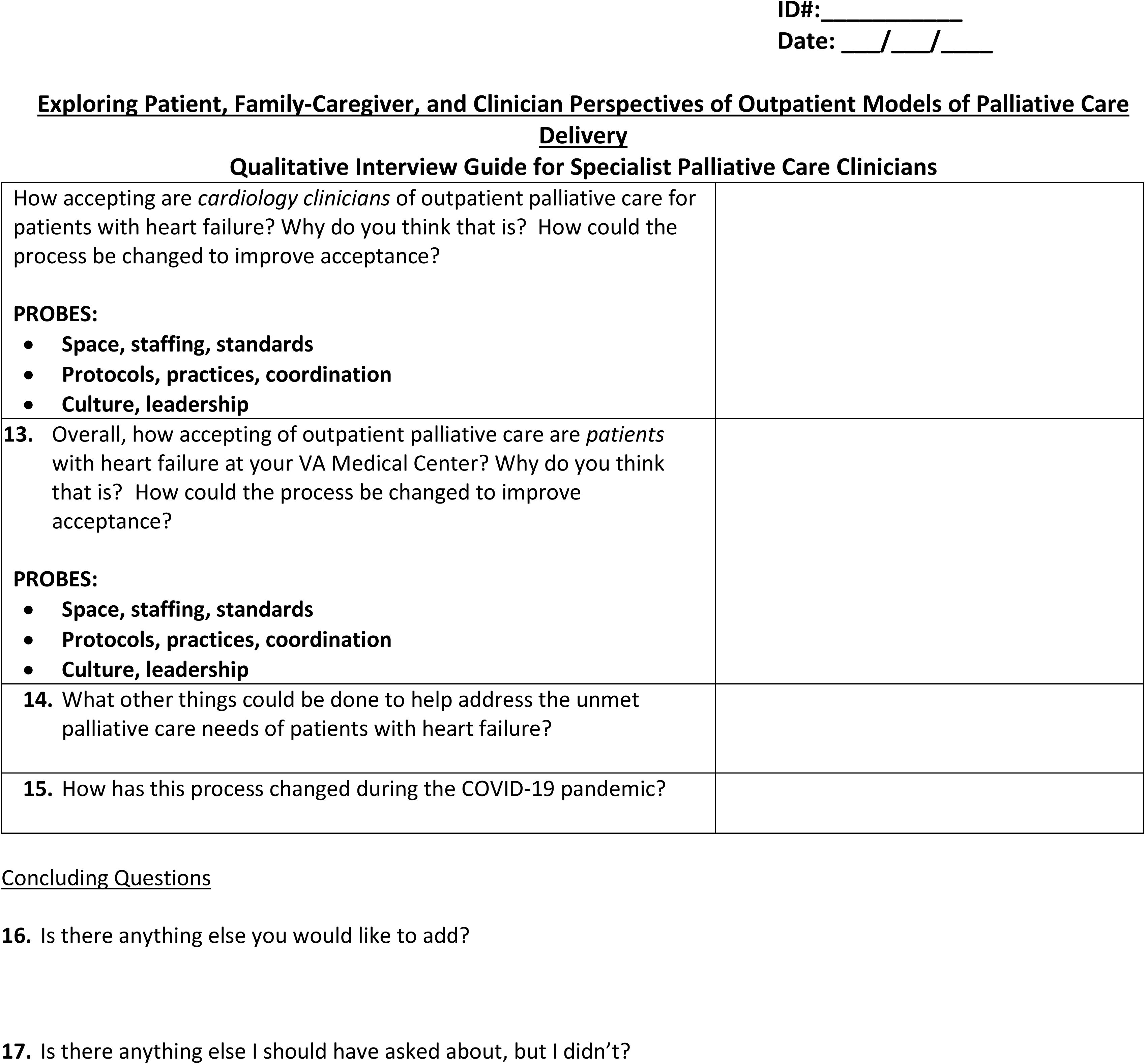

| Grand Tour and General Questions |  |
| --- | --- |
| Question | Notes |
| 1. What are your general thoughts about the value of palliative care for patients with heart failure? What about specifically within outpatient settings? |  |
| 2. What do you consider to be a successful outpatient palliative care consultation for patients with heart failure? |  |
| 3. Tell me about a successful patient encounter with outpatient palliative care. |  |
| 4. Walk me through how patients with heart failure are referred to and receive outpatient palliative care at your VA Medical Center. How did this practice start? Who was the champion of this process? What about before COVID? |  |

| **Components of Outpatient Specialist Palliative Care (i.e. What the Palliative Care Team *does*)** |  |
| --- | --- |
| **Question** | **Notes** |
| **5.** What are key things you address during your visits with patients with heart failure in the outpatient setting? How might this differ from other patient populations?<br>**PROBES:**<br>• **How does this differ from inpatient consults?** |  |
| **6.** What do you think is the most helpful intervention <u>your team</u> provides in the outpatient setting to patients with heart failure and their family-caregivers? |  |
| **7.** What do you think is the least helpful intervention <u>your team</u> provides in the outpatient setting to patients with heart failure and their family-caregivers? |  |
| **8.** What are the characteristics (e.g. clinical factors related to heart failure diagnosis, social factors, others) of patients that are ideally referred for outpatient palliative care?<br>**PROBES:**<br>• **Do these patient characteristics differ from those patients that are typically referred?**<br>• **What are the reasons to refer or to not refer?**<br>• **How do these patients different from inpatient referrals?** |  |

| **Implementation Strategies (i.e. the *characteristics*)** |  |
| --- | --- |
| Question | Notes |
| **9.** What do you think works well in the process of providing palliative care to heart failure patients within outpatient settings? Tell me about some of the specifics, for example the space, staffing, and standards of outpatient palliative care delivery or the protocols, practices, and coordination.<br>**PROBES**<br>• **Culture, leadership** |  |
| **10.** What do you think does not work well in the process of providing palliative care to patients with heart failure within outpatient settings? What could be done to improve this process?<br>**PROBES:**<br>For example, tell me about things related to:<br>• **Space, staffing, standards**<br>• **Protocols, practices, coordination**<br>• **Culture, leadership** |  |
| **11.** What do you think <u>your team</u> could do to improve and/or sustain the process of providing palliative care to patients with heart failure within outpatient settings?<br>**PROBES:**<br>For example, tell me about things related to:<br>• **Space, staffing, standards**<br>• **Protocols, practices, coordination efforts**<br>• **Culture, leadership** |  |
| **12.** What is the role of VAMC and specialty care culture and leadership in the process of providing palliative care to patients with heart failure within outpatient settings? |  |

**Qualitative Interview Guide for Specialist Palliative Care Clinicians**
|  |
| --- |
| How accepting are *cardiology clinicians* of outpatient palliative care for patients with heart failure? Why do you think that is? How could the process be changed to improve acceptance?<br>**PROBES:**<br>• Space, staffing, standards<br>• Protocols, practices, coordination<br>• Culture, leadership |
| **13.** Overall, how accepting of outpatient palliative care are *patients* with heart failure at your VA Medical Center? Why do you think that is? How could the process be changed to improve acceptance?<br>**PROBES:**<br>• Space, staffing, standards<br>• Protocols, practices, coordination<br>• Culture, leadership |
| **14.** What other things could be done to help address the unmet palliative care needs of patients with heart failure? |
| **15.** How has this process changed during the COVID-19 pandemic? |

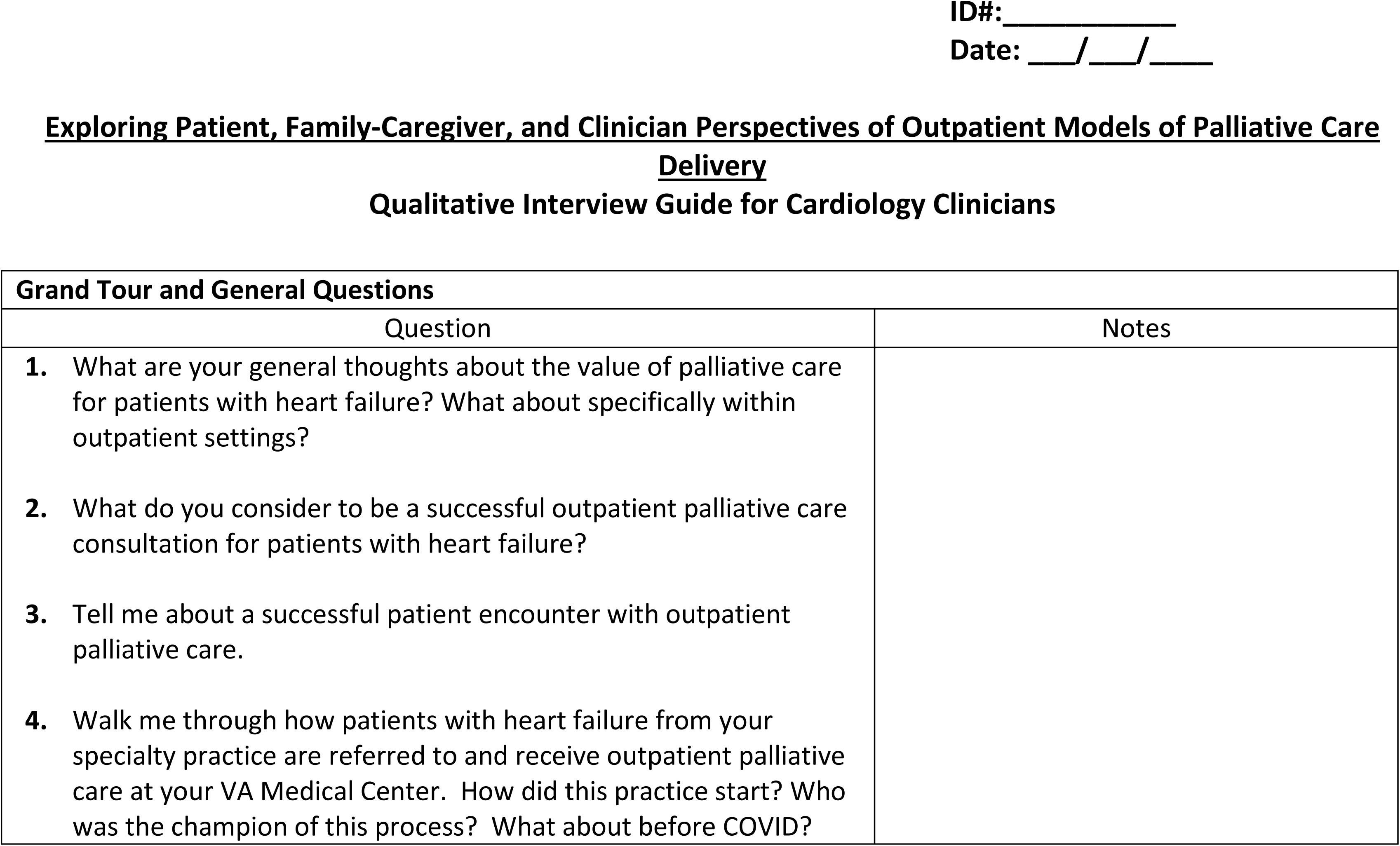
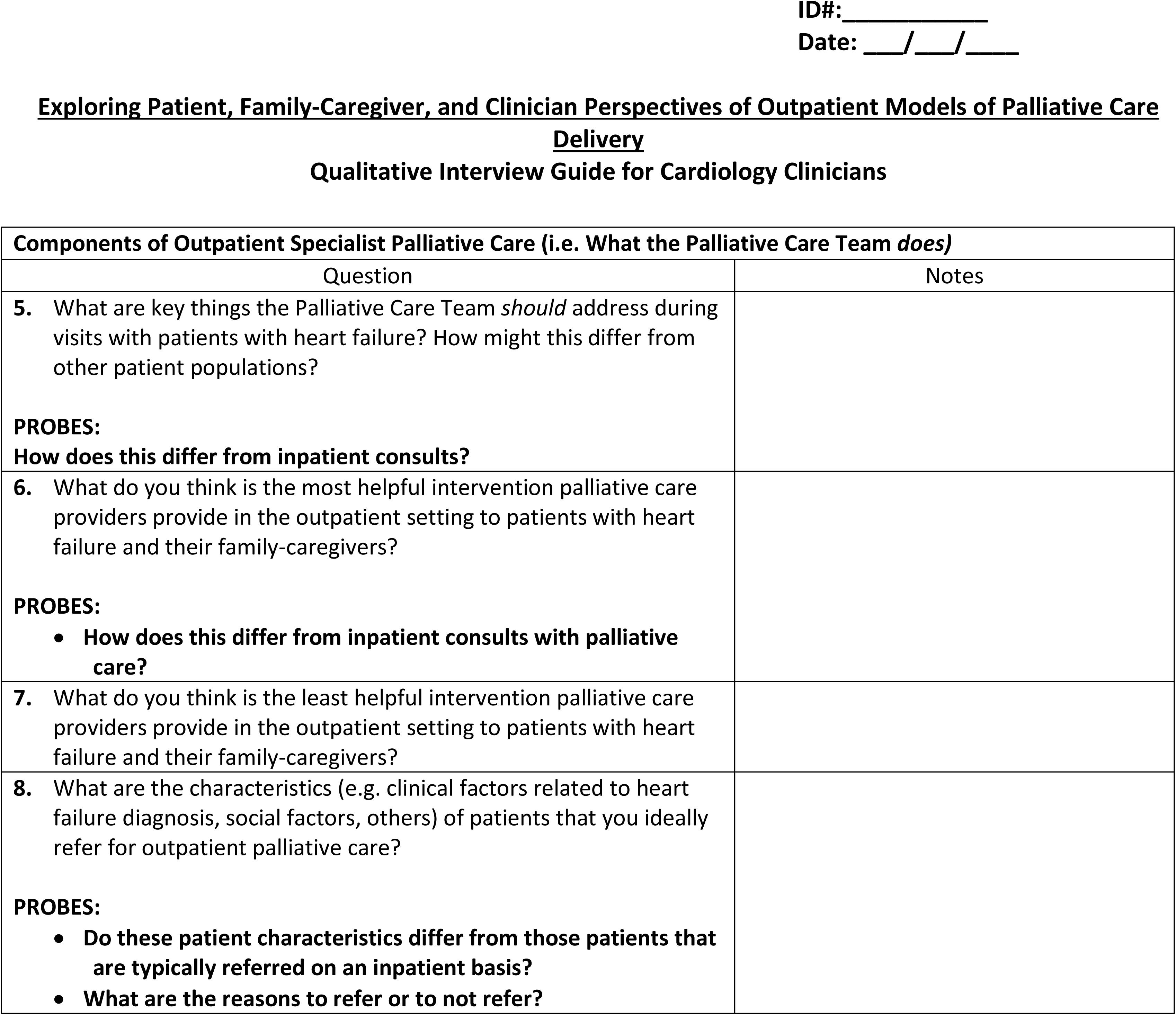
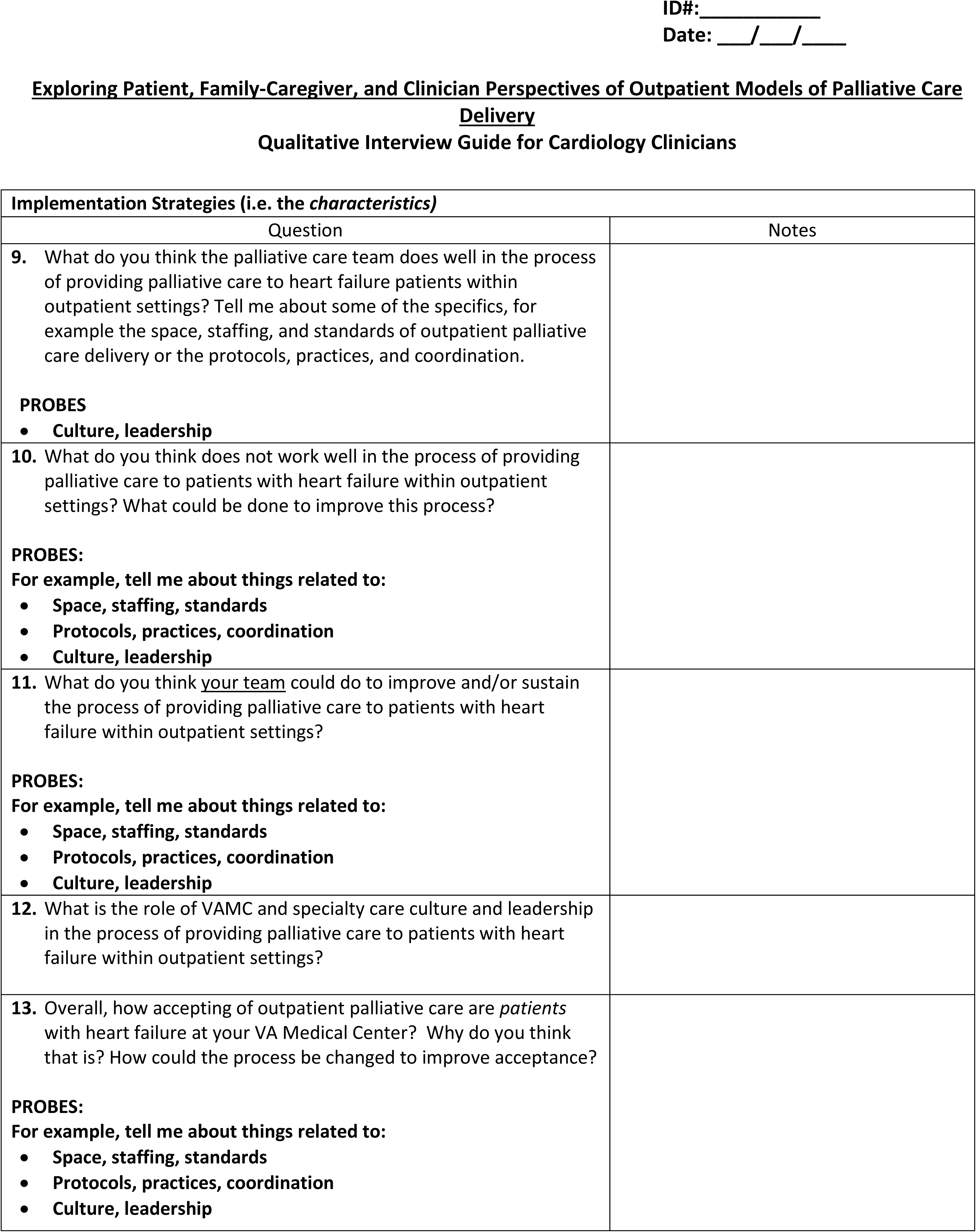
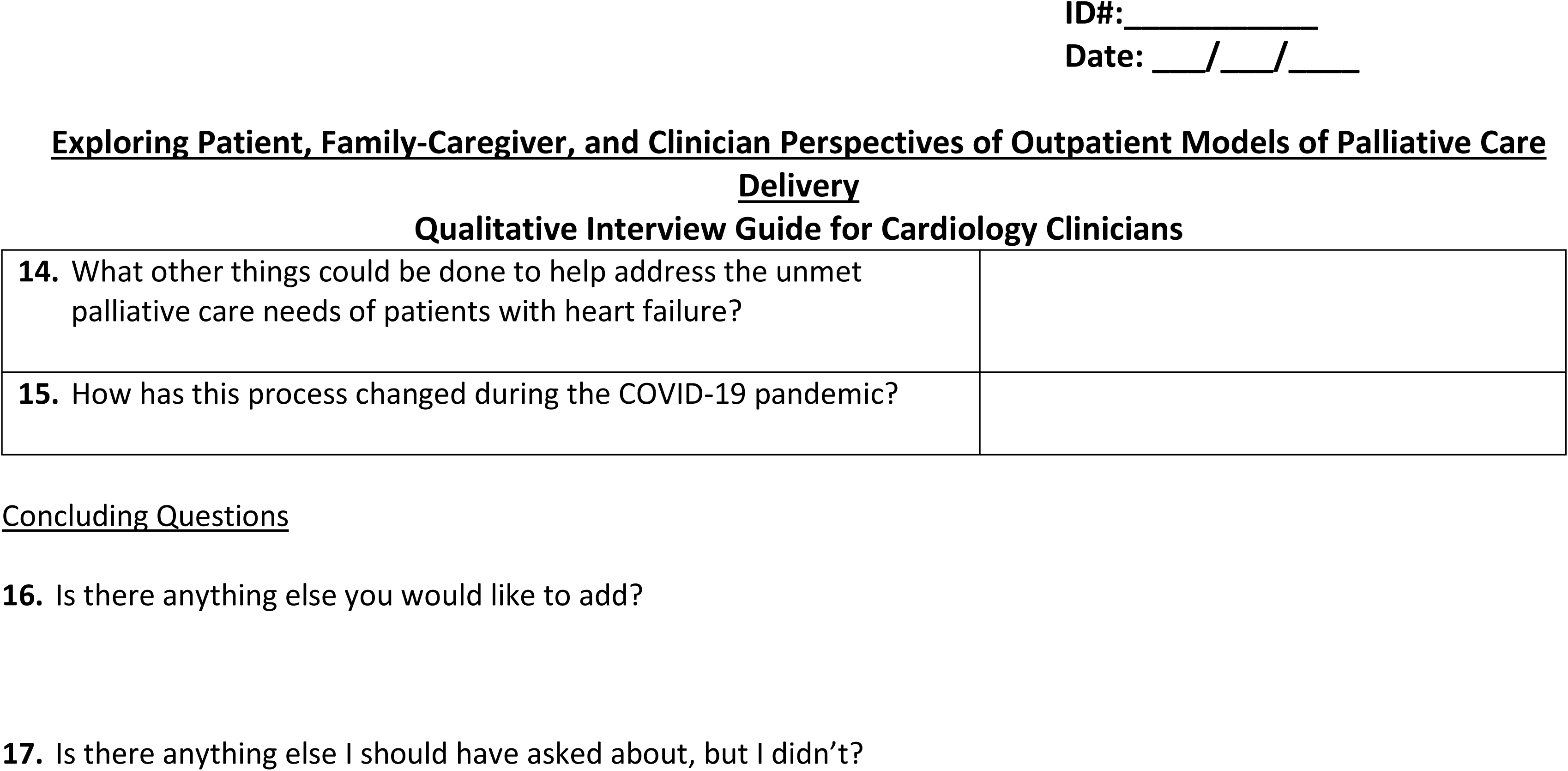

| Grand Tour and General Questions |  |
| --- | --- |
| Question | Notes |
| 1. What are your general thoughts about the value of palliative care for patients with heart failure? What about specifically within outpatient settings? |  |
| 2. What do you consider to be a successful outpatient palliative care consultation for patients with heart failure? |  |
| 3. Tell me about a successful patient encounter with outpatient palliative care. |  |
| 4. Walk me through how patients with heart failure from your specialty practice are referred to and receive outpatient palliative care at your VA Medical Center. How did this practice start? Who was the champion of this process? What about before COVID? |  |

| **Components of Outpatient Specialist Palliative Care (i.e. What the Palliative Care Team *does*)** |  |
| --- | --- |
| Question | Notes |
| **5.** What are key things the Palliative Care Team *should* address during visits with patients with heart failure? How might this differ from other patient populations?<br>**PROBES:**<br>**How does this differ from inpatient consults?** |  |
| **6.** What do you think is the most helpful intervention palliative care providers provide in the outpatient setting to patients with heart failure and their family-caregivers?<br>**PROBES:**<br>• **How does this differ from inpatient consults with palliative care?** |  |
| **7.** What do you think is the least helpful intervention palliative care providers provide in the outpatient setting to patients with heart failure and their family-caregivers? |  |
| **8.** What are the characteristics (e.g. clinical factors related to heart failure diagnosis, social factors, others) of patients that you ideally refer for outpatient palliative care?<br>**PROBES:**<br>• **Do these patient characteristics differ from those patients that are typically referred on an inpatient basis?**<br>• **What are the reasons to refer or to not refer?** |  |

| Implementation Strategies (i.e. the *characteristics*) |  |
| --- | --- |
| Question | Notes |
| 9. What do you think the palliative care team does well in the process of providing palliative care to heart failure patients within outpatient settings? Tell me about some of the specifics, for example the space, staffing, and standards of outpatient palliative care delivery or the protocols, practices, and coordination.<br>**PROBES**<br>• Culture, leadership |  |
| 10. What do you think does not work well in the process of providing palliative care to patients with heart failure within outpatient settings? What could be done to improve this process?<br>**PROBES:**<br>For example, tell me about things related to:<br>• Space, staffing, standards<br>• Protocols, practices, coordination<br>• Culture, leadership |  |
| 11. What do you think <u>your team</u> could do to improve and/or sustain the process of providing palliative care to patients with heart failure within outpatient settings?<br>**PROBES:**<br>For example, tell me about things related to:<br>• Space, staffing, standards<br>• Protocols, practices, coordination<br>• Culture, leadership |  |
| 12. What is the role of VAMC and specialty care culture and leadership in the process of providing palliative care to patients with heart failure within outpatient settings? |  |
| 13. Overall, how accepting of outpatient palliative care are *patients* with heart failure at your VA Medical Center? Why do you think that is? How could the process be changed to improve acceptance?<br>**PROBES:**<br>For example, tell me about things related to:<br>• Space, staffing, standards<br>• Protocols, practices, coordination<br>• Culture, leadership |  |

**Qualitative Interview Guide for Cardiology Clinicians**
|  |
| --- |
| **14.** What other things could be done to help address the unmet palliative care needs of patients with heart failure? |
| **15.** How has this process changed during the COVID-19 pandemic? |

